# Factors associated with hospitalization and critical illness among 4,103 patients with Covid-19 disease in New York City

**DOI:** 10.1101/2020.04.08.20057794

**Authors:** Christopher M. Petrilli, Simon A. Jones, Jie Yang, Harish Rajagopalan, Luke O’Donnell, Yelena Chernyak, Katie A. Tobin, Robert J. Cerfolio, Fritz Francois, Leora I. Horwitz

## Abstract

**Background:** Little is known about factors associated with hospitalization and critical illness in Covid-19 positive patients.

**Methods:** We conducted a cross-sectional analysis of all patients with laboratory-confirmed Covid-19 treated at an academic health system in New York City between March 1, 2020 and April 2, 2020, with follow up through April 7, 2020. Primary outcomes were hospitalization and critical illness (intensive care, mechanical ventilation, hospice and/or death). We conducted multivariable logistic regression to identify risk factors for adverse outcomes, and maximum information gain decision tree classifications to identify key splitters.

**Results:** Among 4,103 Covid-19 patients, 1,999 (48.7%) were hospitalized, of whom 981/1,999 (49.1%) have been discharged, and 292/1,999 (14.6%) have died or been discharged to hospice. Of 445 patients requiring mechanical ventilation, 162/445 (36.4%) have died. Strongest hospitalization risks were age ≥75 years (OR 66.8, 95% CI, 44.7-102.6), age 65-74 (OR 10.9, 95% CI, 8.35-14.34), BMI>40 (OR 6.2, 95% CI, 4.2-9.3), and heart failure (OR 4.3 95% CI, 1.9-11.2). Strongest critical illness risks were admission oxygen saturation <88% (OR 6.99, 95% CI 4.5-11.0), d-dimer>2500 (OR 6.9, 95% CI, 3.2-15.2), ferritin >2500 (OR 6.9, 95% CI, 3.2-15.2), and C-reactive protein (CRP) >200 (OR 5.78, 95% CI, 2.6-13.8). In the decision tree for admission, the most important features were age >65 and obesity; for critical illness, the most important was SpO2<88, followed by procalcitonin >0.5, troponin <0.1 (protective), age >64 and CRP>200.

**Conclusions:** Age and comorbidities are powerful predictors of hospitalization; however, admission oxygen impairment and markers of inflammation are most strongly associated with critical illness.

## Background

The first announcement of a cluster of novel pneumonia-like illness was made on December 31, 2019 by China. Since then, the causative organism, SARS-Cov-2, has spread to cause a global pandemic that to date has infected over a million people and directly resulted in over 75,000 deaths.

While several reports from China,^1^ Italy,^2,3^ and most recently the United States Centers for Disease Control and Prevention^4,5^ have described characteristics of patients with Covid-19, the disease caused by SARS-Cov-2, little is understood about factors associated with hospital admission and with severe disease. Studies to date have included few patients with severe outcomes,^1,6-10^ or have not compared those to patients with less virulent disease,^11-13^ making it difficult to assess characteristics associated with poor outcomes. No studies to date have conducted multivariable regression to help identify the strongest risk factors. Moreover, very few studies to date are from the United States. Differences in population demographics and behaviors may limit generalizability of those characteristics to patients in the United States, which currently has the most Covid-19 cases in the world.

Understanding which patients are most at risk for hospitalization is crucial for many reasons. It can assist emergency providers in making triage decisions and ambulatory clinicians in identifying patients who would most benefit from early treatment once available. It can help inform policymakers about highest risk populations, who may need particular protection in policy determinations. Finally, it can help epidemiologists to improve the accuracy of projections about likely need for hospital beds and staffing needs in a region given its demographic characteristics.^14^

For similar reasons, is also important to understand the risk of critical illness among those hospitalized. Clinicians need this information both to identify patients at greatest risk of deterioration, and to inform decision-making about hospital discharge. Policymakers and epidemiologists need this information to project likely need for intensive care unit capacity, ventilators and associated staffing. It would also help improve the reliability of future mortality rate estimation.

New York City is now the epicenter of the Covid-19 outbreak in the United States, with over 68,000 known cases in the city and over 2,700 deaths as of April 6: more than anywhere else in the country.^15^ In this report, the largest case series from the United States to date, we describe characteristics of Covid-19 patients treated at a large quaternary academic health system in New York City and Long Island, and the association of these characteristics with adverse outcomes.

## Methods

### Study setting

The study was conducted at NYU Langone Health, which includes over 260 outpatient office sites and four acute care hospitals (two in Manhattan, one in Brooklyn, one in Long Island) ranging from a quaternary care hospital to a safety net institution. As the epidemic evolved, the health system added intensive care unit beds and inpatient capacity, resulting in approximately 394 ICU beds and 1,297 non-ICU beds at time of writing.

### Data sources

We obtained data from the electronic health record (Epic Systems, Verona, WI), which is an integrated EHR including all inpatient and outpatient visits in the health system, beginning on March 1, 2020 and ending on April 2, 2020. Follow up was complete through April 7, 2020.

A confirmed case of Covid-19 was defined as a positive result on real-time reverse-transcriptase-polymerase-chain-reaction (RT-PCR) assay of nasopharyngeal or oropharyngeal swab specimens. Initial tests were conducted by the New York City Department of Health and Mental Hygiene; as of March 16, tests were conducted in our clinical laboratory using the Roche SARS-CoV2 assay in the Cobas 6800 instruments through emergency use authorization (EUA) granted by the FDA. On March 31 we added testing using the SARS-CoV2 Xpert Xpress assay in the Cepheid GeneXpert instruments also under EUA by FDA. The targets amplified by these assays are the ORF1/a and E in the Roche Cobas assay and N2 and E genes in the Cepheid XpertXpress. Since March 16, only pharyngeal samples were collected and tested.

Testing was performed for patients presenting to the emergency department with any complaint consistent with Covid-19, including fever, cough, shortness of breath, fatigue, gastrointestinal complaints, syncope, known exposure to a Covid-19 positive patient, or clinician concern. In addition, ambulatory testing was available by appointment with clinician’s referral until March 26, 2020, when New York State recommended restricting testing of patients with mild or moderate illness. Outpatient testing of symptomatic or concerned employees has remained available throughout the study period. Repeat testing of negative specimens was conducted at clinician discretion. If testing was repeated and discordant (i.e. negative test followed by a positive test), we used the positive result.

### Main outcomes

We assessed two primary outcomes: inpatient hospitalization and critical illness, defined as a composite of care in the intensive care unit, use of mechanical ventilation, discharge to hospice, or death. For patients with multiple visits, the most severe outcome was assigned. For instance, patients who did not need hospitalization at time of initial testing but were later hospitalized were assigned to the hospitalization group. Similarly, patients who were initially admitted and discharged and then readmitted requiring invasive ventilation were assigned to the critical illness group.

### Predictors

We obtained from the electronic health record the following variables: age at time of testing, sex, race as reported by the patient (aggregated into white, African American, Asian, other and unknown), ethnicity as reported by the patient (Hispanic or non-Hispanic), any past cardiac history (as defined by a history of hypertension, hyperlipidemia, coronary artery disease or heart failure), any past pulmonary disease (as defined by chronic obstructive pulmonary disease or asthma), malignancy (excluding non-melanoma skin malignancy), diabetes, and obesity (defined by body mass index). We also obtained vital signs and first set of laboratory results where available. For multivariable modeling, we bucketed vital sign and laboratory results into categories by degree of abnormality based on clinical judgment because of non-linear associations with outcome. In the hospitalization analysis, we included only patient demographics and past history, since 1,642/2,104 (78%) patients who were not admitted were evaluated in ambulatory testing centers and did not have vitals or laboratory studies collected. For the critical illness analysis, we included the above predictors and added temperature and oxygen saturation on presentation, as well as the first result of c-reactive protein, d-dimer, ferritin, procalcitonin, and troponin when obtained. We selected these predictors based on prior published literature and our clinical experience with Covid-19 patients.

### Statistical analysis

We used descriptive statistics to characterize each cohort of patients: those not hospitalized, all those hospitalized, those discharged to home, and those with critical illness (care in intensive care unit, mechanical ventilation, discharge to hospice, or death). We then fitted multivariable logistic regression models with admission and with critical illness as the outcomes to identify factors associated with those outcomes. We included all selected predictors based on *a priori* clinical significance after testing for collinearity using the variance inflation factor (VIF) and ensuring none had VIF>2.^16^ For the admission model, we included all patients testing positive. For the critical illness model, we included only patients who had been discharged alive or had suffered severe complications, omitting patients still hospitalized, for whom final outcome was not yet determined. We excluded from this model four patients who expired in the emergency department before any information including vital signs could be collected. We obtained odds ratios from the models and bootstrapped confidence intervals for the odds ratios using the approach of Venables & Ripley,^17^ since assuming normality of the maximum likelihood estimate to estimate confidence intervals can lead to biased estimates.^18^

Finally, we constructed maximum information gain decision tree classifications for both hospital admission and severe complication to identify the variables that best classified patients into different outcome cohorts. For a given population, the decision tree classification method splits the population into two groups using one feature at a time, starting with the feature that maximizes the split between groups relative to the outcome in question.^19,20^ Subsequent splits reevaluate each split subgroup for the next best feature. The final population in each end node has similar characteristics and outcomes. We used the decision tree classifier from Python 3.7.4 scikit-learn library. We chose to maximize information gain (which minimizes entropy) for each branch split in the classification tasks. We also pruned the trees to prevent overfitting by limiting the maximum depth, minimum samples in a leaf, and minimum sample splits. For both models, we split the data into a training set (80%) and hold-out set (20%). For the admission model, we ran 48 iterations to achieve optimal parameters. For the complications model, we ran 64 iterations.

The logistic regression models were conducted with R, version 3.6.3, and the decision trees with Python, version 3.7.4. All analyses used 2-sided statistical tests and we considered a p value < 0.05 to be statistically significant without adjustment for multiple testing.

This study was approved by the NYU Grossman School of Medicine Institutional Review Board, which granted both a waiver of informed consent, and a waiver of the Health Information Portability and Privacy Act.

## Results

During the study period, the health system tested 7,719 patients for Covid-19, of whom 4,103 (48.7%) were positive. Of patients testing positive, 2,104 (51.3%) were treated as outpatients, and 1,999 (48.7%) were admitted to the hospital. Among those admitted to the hospital, 1,582 (79.1%) have experienced a study outcome, among which 932/1,582 (58.9%) were discharged without complication and 650/1,582 (41.1%) experienced critical illness, including 292/1,582 (18.5%) who have been discharged to hospice or died.

Among the 650 patients with critical illness, 445/650 (68.5%) required mechanical ventilation, 89/650 (13.7%) required intensive care without mechanical ventilation, and 116/650 (17.8%) were discharged to hospice or died without either intensive care or mechanical ventilation. Final outcomes to date for each subgroup are shown in **Figure 1**.

**Figure 1:**
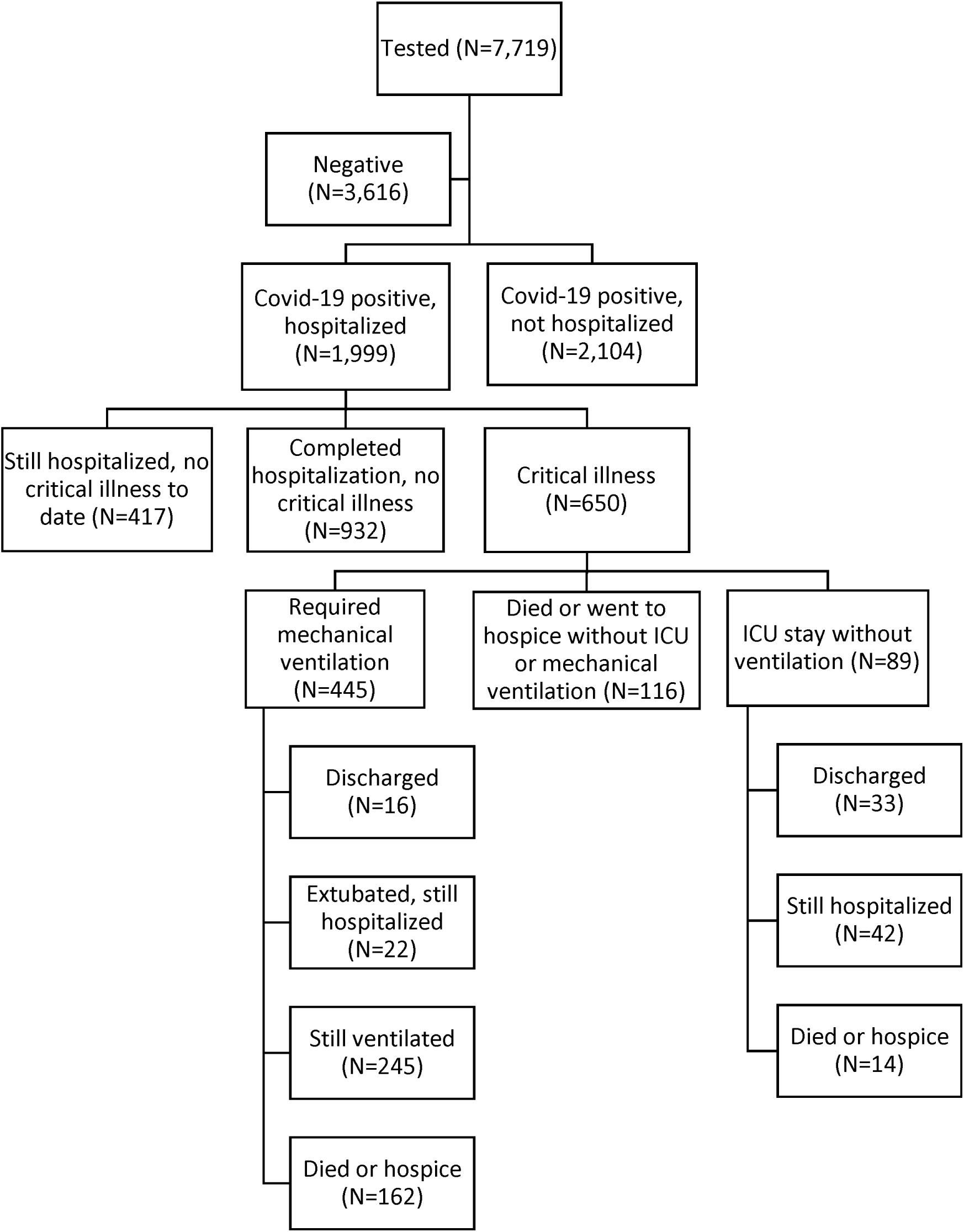
Flow diagram of included patients.

### Characteristics of study population

The median age of the Covid-19 positive study population was 52 years (interquartile range, 36 to 65), and 2,072 (50.5%) were male. A total of 614 (15.0%) had diabetes, 1,100 (26.8%) obesity, and 1,235 (30.1%) cardiovascular disease. Among hospitalized patients, the median length of stay among those with final discharge disposition (discharged alive or died) was 4.8 days (interquartile range, 3.3 to 7.6). Median days of follow up for those still hospitalized with critical illness was 11.4 (IQR 8.4 to 15.4).

Hospitalized patients were more likely to be male (62.6% vs 39.0%) and had substantially more comorbidities than non-hospitalized patients, particularly with regard to cardiovascular disease (44.6% vs. 16.4%), diabetes (31.8% vs 5.4%) and obesity (39.8% vs. 14.5%) (**Table 1**). Differences in sex and comorbidities between patients experiencing severe deterioration and those who did not were much smaller. Among these patients, differences in clinical presentation and laboratory results were more prominent. Patients experiencing severe deterioration were more likely to present with hypoxia (initial O2 saturation 25^th^ percentile 86% versus 93%), and had higher initial levels of c-reactive protein (median 139 vs 80.8), d-dimer (median 513 vs 306), ferritin (median 980.5 vs 574.5). (**Table 2**).

**Table 1:** Characteristics of tested patients, by hospitalization status.

| Characteristic | Not hospitalized, N=2,104<br>N (%) or median (IQR) | Hospitalized, N=1,999<br>N (%) or median (IQR) |
| --- | --- | --- |
| Age | 41 (31, 54) | 62 (50, 74) |
| Male sex | 821 (39.0) | 1,251 (62.6) |
| Race |  |  |
| White | 887 (42.2) | 925 (46.3) |
| African American | 365 (17.3) | 290 (14.5) |
| Asian | 166 (7.9) | 118 (5.9) |
| Other | 391 (18.6) | 536 (26.8) |
| Multiracial/Unknown | 295 (14.0) | 130 (6.5) |
| Tobacco use |  |  |
| Never or unknown | 1746 (83.0) | 1479 (74.0) |
| Former | 250 (11.9) | 416 (20.8) |
| Current | 108 (5.1) | 104 (5.2) |
| Obesity | 304 (14.4) | 796 (39.8) |
| BMI 30-40 kg/m<sup>2</sup> | 256 (12.2) | 659 (33.0) |
| BMI >40 kg/m<sup>2</sup> | 48 (2.3) | 137 (6.9) |
| Any chronic condition* | 629 (29.9) | 1438 (71.9) |
| Any cardiovascular condition* | 344 (16.3) | 891 (44.6) |
| Hyperlipidemia | 221 (10.5) | 517 (25.9) |
| Hypertension | 241 (11.5) | 742 (37.1) |
| Coronary artery disease | 38 (1.8) | 197 (9.9) |
| Heart failure | 7 (0.3) | 124 (6.2) |
| Diabetes | 111 (5.3) | 503 (25.2) |
| Asthma or chronic obstructive pulmonary disorder | 106 (5.0) | 206 (10.3) |
| Chronic kidney disease | 20 (1.0) | 195 (9.8) |
| Malignancy | 42 (2.0) | 143 (7.2) |
| Temperature at presentation, degrees Celsius | 37.3 (36.9 – 37.8) | 37.5 (36.9 – 38.3) |
| Temperature ≥ 38 at presentation | 106 (5.0)** | 669 (33.5) |
| Oxygen saturation at presentation | 97 (96-99) **† | 94 (91-97) ‡ |

\*Not included in multivariable model
\*\*Missing for 76% of non-hospitalized patients
†Measured on supplemental oxygen for 0.3% of patients
‡Measured on supplemental oxygen for 78.9% of patients

**Table 2:** Characteristics of admitted patients, by complication status.

| **Characteristic** | **Discharged, no critical illness, N=932**<br>**N (%) or median (IQR)** | **Critical illness, N=650**<br>**N (%) or median (IQR)** |
| --- | --- | --- |
| Age, years | 58 (46-71) | 67 (56-77) |
| Male sex | 560 (60.1) | 442 (68.0) |
| Race |  |  |
| White | 411 (44.1) | 314 (48.3) |
| African American | 160 (17.2) | 74 (11.4) |
| Asian | 43 (4.6) | 52 (8.0) |
| Other | 264 (28.3) | 161 (24.8) |
| Multiracial/Unknown | 54 (5.8) | 49 (7.5) |
| Tobacco use |  |  |
| Never/unknown | 695 (74.6) | 477 (73.4) |
| Former | 175 (18.8) | 145 (22.3) |
| Current | 62 (6.7) | 28 (4.3) |
| Obesity |  |  |
| BMI < 30 kg/m<sup>2</sup> | 504 (54.1) | 359 (55.2) |
| BMI 30-40 kg/m<sup>2</sup> | 312 (33.5) | 210 (32.3) |
| BMI > 40 kg/m<sup>2</sup> | 66 (7.1) | 50 (7.7) |
| Any chronic condition* | 659 (70.7) | 477 (73.4) |
| Any cardiovascular condition* | 391 (42.0) | 306 (47.1) |
| Hypertension | 320 (34.3) | 257 (39.5) |
| Hyperlipidemia | 225 (24.1) | 173 (26.6) |
| Coronary artery disease | 79 (8.5) | 80 (12.3) |
| Heart failure | 47 (5.0) | 54 (8.3) |
| Diabetes | 213 (22.9) | 176 (27.1) |
| Cancer | 54 (5.8) | 56 (8.6) |
| Asthma or chronic obstructive pulmonary disorder | 91 (9.8) | 71 (10.9) |
| Chronic kidney disease | 81 (8.7) | 75 (11.5) |
| Temperature at presentation, degrees Celsius | 37.5 (36.9 – 38.3) | 37.6 (36.9 – 38.4) |
| Temperature ≥ 38 at presentation | 315 (74.6) | 226 (73.4) |
| Oxygen saturation at presentation | 95 (93-97) † | 92 (86-95) ‡ |
| First absolute lymphocyte count, 10<sup>3</sup>/μl | 0.9 (0.7-1.3) | 0.8 (0.5-1.1) |
| First creatinine, mg/dL | 0.93 (.77-1.2) | 1.06 (.83-1.5) |
| First alanine aminotransferase, units/L* | 32 (22-52) | 35 (24-53) |
| First aspartate aminotransferase, units/L* | 39 (28-57) | 52 (36-78) |
| First C-reactive protein, mg/L | 80.8 (36.8-136) | 139.3 (139.3-201.9) |
| First d-dimer, ng/mL | 306 (201-497) | 513 (305-1087) |

| Characteristic | Discharged, no critical illness, N=932 | Critical illness, N=650 |
| --- | --- | --- |
|  | N (%) or median (IQR) | N (%) or median (IQR) |
| First ferritin, ng/mL | 574.5 (280-1277) | 980.5 (520-1765) |
| First procalcitonin, ng/mL | 0.09 (0.05-0.20) | 0.27 (0.11-0.82) |
| First troponin, ng/mL | 0.015 (0.01-0.09) | 0.06 (0.02-0.09) |
| Length of stay, days* | 4 (2-6) | 6 (3-9) [discharged or died]<br>11 (8-15) [still hospitalized] |

\*Not included in multivariable model
†Measured on supplemental oxygen for 67.7% of patients
‡Measured on supplemental oxygen for 88.2% of patients

### Predictors of hospitalization

In multivariable analysis, the factors most associated with hospitalization were age 75 years or older (OR 66.8, 95% CI, 44.7 to 102.6), age 65-74 (OR 10.9, 95% CI, 8.35 to 14.34), BMI>40 (OR 6.2, 95% CI, 4.2-9.3), and history of heart failure (OR 4.3 95% CI, 1.9-11.2). Full model results are shown in **Table 3**.

**Table 3:** Multivariable regression results, hospitalization.

| Characteristic | N (%) | Odds ratio (95% CI) | p |
| --- | --- | --- | --- |
| Age, years |  |  |  |
| 0-18 | 53 (1.3%) | 4.96 (2.75-8.98) | <0.001 |
| 19-44 | 1501 (36.6%) | Reference | <0.001 |
| 45-54 | 694 (16.9%) | 2.57 (2.06-3.2) | <0.001 |
| 55-64 | 767 (18.7%) | 4.17 (3.35-5.2) | <0.001 |
| 65-74 | 577 (14.1%) | 10.91 (8.35-14.34) | <0.001 |
| ≥75 | 511 (12.5%) | 66.79 (44.73-102.62) | <0.001 |
| Cancer | 185 (4.5%) | 1.24 (0.81-1.93) | 0.329 |
| Chronic kidney disease | 215 (5.2%) | 3.07 (1.78-5.52) | <0.001 |
| Coronary artery disease | 235 (5.7%) | 0.88 (0.57-1.4) | 0.590 |
| Diabetes | 614 (15%) | 2.81 (2.14-3.72) | <0.001 |
| Male | 2072 (50.5%) | 2.8 (2.38-3.3) | <0.001 |
| Heart failure | 131 (3.2%) | 4.29 (1.89-11.18) | 0.001 |
| Hyperlipidemia | 738 (18%) | 0.67 (0.51-0.87) | 0.003 |
| Hypertension | 983 (24%) | 1.23 (0.97-1.57) | 0.094 |
| Obesity |  |  |  |
| BMI <30 kg/m<sup>2</sup> | 3003 (73.2%) | Reference | <0.001 |
| BMI 30-40 kg/m<sup>2</sup> | 915 (22.3%) | 4.26 (3.5-5.2) | <0.001 |
| BMI >40 kg/m<sup>2</sup> | 185 (4.5%) | 6.2 (4.21-9.25) | <0.001 |
| Pulmonary disease | 312 (7.6%) | 1.33 (0.96-1.84) | 0.087 |
| Race |  |  |  |
| White | 1812 (44.2%) | Reference | <0.001 |
| African American | 655 (16%) | 0.88 (0.69-1.11) | 0.280 |
| Asian | 284 (6.9%) | 1.44 (1.04-1.98) | 0.026 |
| Other/Multiracial | 927 (22.6%) | 1.99 (1.62-2.45) | <0.001 |
| Unknown | 425 (10.4%) | 0.9 (0.67-1.21) | 0.501 |
| Tobacco use (current or former) | 878 (21.4%) | 0.71 (0.57-0.87) | 0.001 |

The factors most associated with critical illness were admission oxygen saturation <88% (OR 6.99, 95% CI 4.5 to 11.0), first d-dimer>2500 (OR 6.9, 95% CI, 3.2 to 15.2), first ferritin >2500 (OR 6.9, 95% CI, 3.2-15.2), and first C-reactive protein (CRP) >200 (OR 5.78, 95% CI, 2.6 to 13.8). Age 0-18 had a high OR of 6.3 (95% CI, 2.4 to 16.1), but this group included only 28 patients, of whom 19 were newborns; most of the critically ill were > 16 years. There were no deaths in this age group. See **Table 4** for full model results.

**Table 4:** Multivariable regression results, critical illness.

| Characteristic | N (%) | Odds ratio (95% CI) | p |
| --- | --- | --- | --- |
| Age, years |  |  |  |
| 0-18 | 28 (1.8%) | 6.25 (2.39-16.10) | <0.001 |
| 19-44 | 267 (16.9%) | Reference |  |
| 45-54 | 248 (15.7%) | 0.71 (0.44-1.15) | 0.165 |
| 55-64 | 328 (20.7%) | 1.25 (0.81-1.94) | 0.309 |
| 65-74 | 343 (21.7%) | 1.88 (1.20-2.95) | 0.006 |
| ≥75 | 368 (23.3%) | 2.57 (1.62-4.11) | <0.001 |
| Cancer | 110 (7.0%) | 1.14 (0.67-1.91) | 0.632 |
| Chronic kidney disease | 156 (9.9%) | 0.51 (0.29-0.89) | 0.018 |
| Coronary artery disease | 159 (10.1%) | 0.89 (0.55-1.41) | 0.614 |
| Diabetes | 389 (24.6%) | 1.14 (0.83-1.58) | 0.411 |
| Male | 1002 (63.3%) | 0.99 (0.74-1.33) | 0.953 |
| Heart failure | 101 (6.4%) | 1.31 (0.73-2.34) | 0.358 |
| Hyperlipidemia | 398 (25.2%) | 0.96 (0.68-1.37) | 0.841 |
| Hypertension | 577 (36.5%) | 0.95 (0.68-1.33) | 0.781 |
| Obesity |  |  |  |
| BMI <30 kg/m<sup>2</sup> | 944 (59.7%) | Reference |  |
| BMI 30-40 kg/m<sup>2</sup> | 522 (33.0%) | 1.38 (1.03-1.85) | 0.029 |
| BMI >40 kg/m<sup>2</sup> | 116 (7.3%) | 1.73 (1.03-2.90) | 0.038 |
| Pulmonary disease | 162 (10.2%) | 1.21 (0.79-1.86) | 0.380 |
| Race |  |  |  |
| White | 725 (45.8%) | Reference |  |
| African American | 234 (14.8%) | 0.58 (0.39-0.87) | 0.009 |
| Asian | 95 (6.0%) | 1.91 (1.09-3.37) | 0.024 |
| Other/Multiracial | 425 (26.9%) | 0.95 (0.69-1.30) | 0.751 |
| Unknown | 103 (6.5%) | 1.11 (0.65-1.88) | 0.695 |
| Tobacco use (current or former) | 410 (25.9%) | 0.89 (0.65-1.21) | 0.452 |
| First C reactive protein, mg/L |  |  |  |
| 0-15 | 101 (6.4%) | Reference |  |
| >15-100 | 531 (33.6%) | 4.88 (2.38-11.00) | <0.001 |
| >100-200 | 494 (31.2%) | 2.86 (1.42-6.36) | 0.006 |
| >200 | 228 (14.4%) | 5.78 (2.62-13.76) | <0.001 |
| Missing | 228 (14.4%) | 2.93 (1.15-7.86) | 0.027 |
| First creatinine, mg/dL |  |  |  |
| 0-1.2 | 947 (59.9%) | Reference |  |
| >1.2-2 | 382 (24.1%) | 1.39 (1.01-1.92) | 0.042 |
| >2 | 146 (9.2%) | 1.73 (0.94-3.19) | 0.077 |
| Missing | 107 (6.8%) | 0.97 (0.36-2.55) | 0.956 |
| First d-dimer, ng/mL |  |  |  |
| 0-250 | 319 (20.2%) | Reference |  |
| >250-500 | 421 (26.6%) | 1.64 (1.13-2.41) | 0.010 |
| >500-1000 | 252 (15.9%) | 2.41 (1.58-3.70) | <0.001 |
| >1000-2500 | 82 (5.2%) | 4.88 (2.56-9.51) | <0.001 |
| >2500 | 67 (4.2%) | 6.85 (3.24-15.20) | <0.001 |
| Missing | 441 (27.9%) | 1.36 (0.86-2.13) | 0.188 |
| First ferritin, ng/mL |  |  |  |
| 0-300 | 265 (16.8%) | Reference |  |
| >300-500 | 191 (12.1%) | 1.48 (0.90-2.43) | 0.121 |
| >500-1000 | 331 (20.9%) | 1.77 (1.14-2.75) | 0.011 |
| >1000-2500 | 372 (23.5%) | 1.63 (1.05-2.56) | 0.032 |
| >2500 | 136 (8.6%) | 1.64 (0.93-2.90) | 0.090 |
| Missing | 287 (18.1%) | 2.08 (1.11-3.91) | 0.023 |
| First lymphocyte count, 10<sup>3</sup>/μl |  |  |  |
| >1.2 | 360 (22.8%) | Reference |  |
| >0.8-1.2 | 388 (24.5%) | 0.96 (0.66-1.41) | 0.843 |
| 0.4-0.8 | 596 (37.7%) | 1.12 (0.78-1.59) | 0.541 |
| <0.4 | 184 (11.6%) | 2.15 (1.34-3.47) | 0.002 |
| Missing | 54 (3.4%) | 2.64 (0.96-7.57) | 0.063 |
| First procalcitonin, ng/mL |  |  |  |
| 0-0.5 | 1065 (67.3%) | Reference |  |
| >0.5 | 280 (17.7%) | 1.84 (1.26-2.68) | 0.002 |
| Missing | 237 (15.0%) | 0.44 (0.24-0.79) | 0.007 |
| Oxygen saturation on presentation, % |  |  |  |
| SpO2 >92 | 1029 (65.0%) | Reference |  |
| SpO2 88-92 | 336 (21.2%) | 1.48 (1.09-2.01) | 0.012 |
| SpO2 <88 | 215 (13.6%) | 6.99 (4.54-11.02) | <0.001 |
| Missing | 2 (0.1%) |  |  |
| Temperature on presentation, degrees Celsius |  |  |  |
| <38 | 438 (27.7%) | Reference |  |
| 38-39 | 953 (60.2%) | 0.89 (0.66-1.20) | 0.457 |
| >39 | 191 (12.1%) | 1.49 (0.96-2.31) | 0.073 |
| First troponin, ng/mL |  |  |  |
| <0.1 | 1142 (72.2%) | Reference |  |
| 0.1-1 | 148 (9.4%) | 2.56 (1.59-4.20) | 0.001 |
| >1 | 37 (2.3%) | 11.00 (2.94-57.90) | 0.741 |
| Missing | 255 (16.1%) | 0.92 (0.55-1.52) | <0.001 |

In a maximum information gain decision tree for admission, the most important feature at the top-level branch point was age >65, followed by obesity; for critical illness, the top branch point was SpO2<88, followed by procalcitonin >0.5, troponin <0.1 (protective), age >64 and CRP>200. (See **Figure 2A and Figure 2B**)

**Figure 2A:**
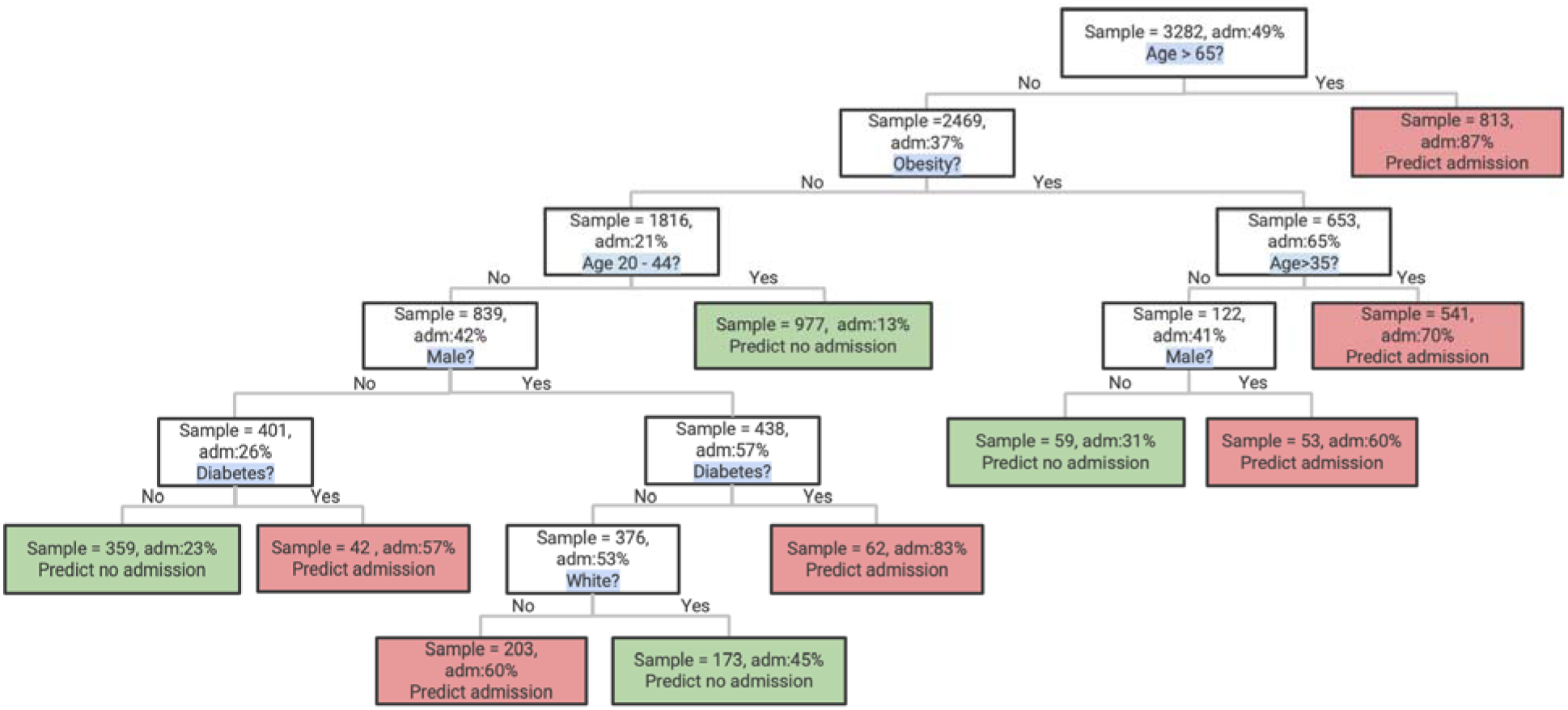
Maximum likelihood classification tree for hospitalization.

**Figure 2B:**
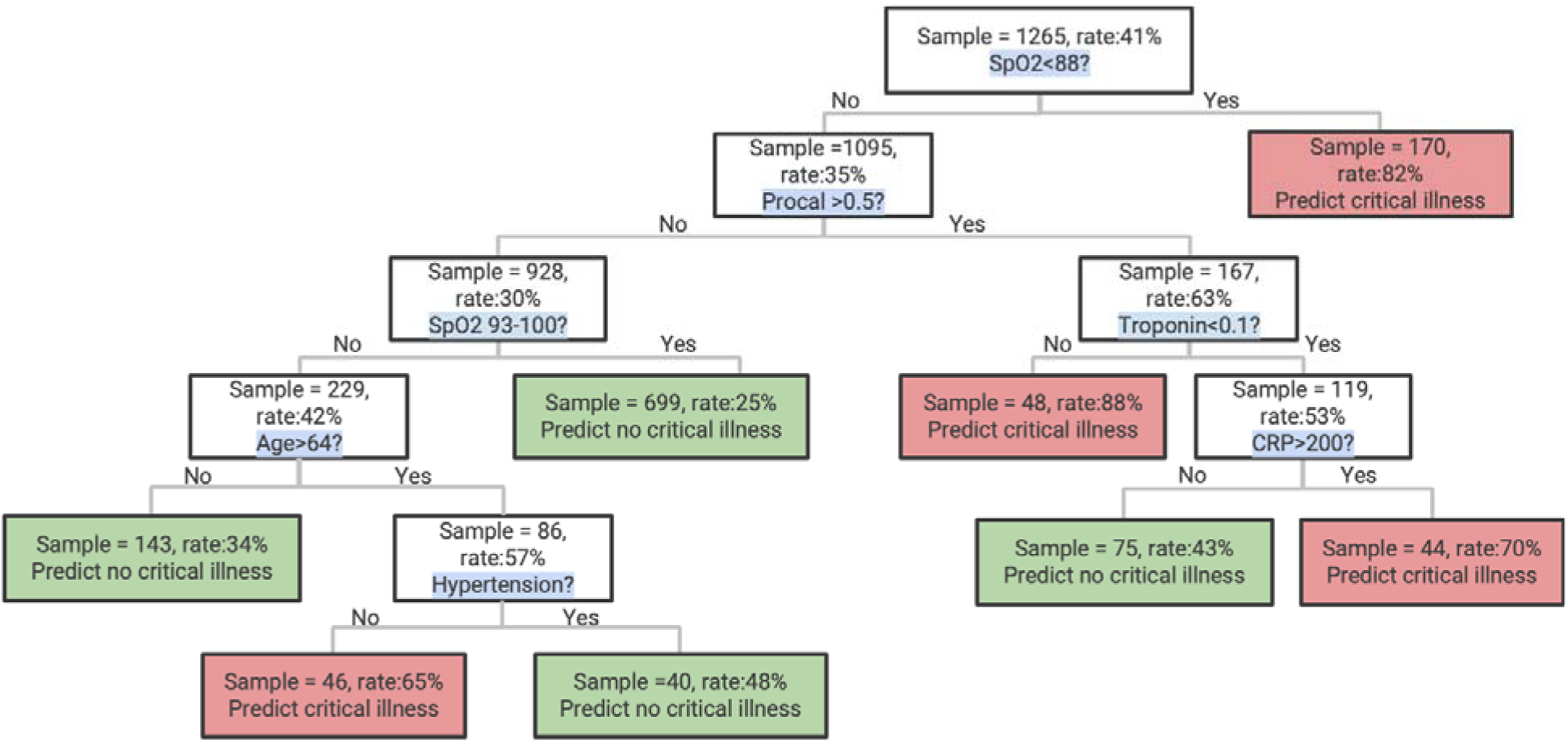
Maximum likelihood classification tree for critical illness.

## Discussion

In this report, we describe characteristics of 4,103 patients with laboratory-confirmed Covid-19 disease in New York City, of whom 1,999 required hospital admission and 650 required intensive care, mechanical ventilation, were discharged to hospice and/or died. We find particularly strong associations of older age, obesity, heart failure and chronic kidney disease with hospitalization risk, with much less influence of race, smoking status, chronic pulmonary disease and other forms of heart disease. Moreover, we also noted the importance of hypoxia despite supplemental oxygen and early elevations in inflammatory markers (especially d-dimer and c-reactive protein) in distinguishing among patients who go on to develop critical illness and those who do not. In the hospitalized population, measures of inflammation were much more important than demographic characteristics and comorbidities.

The largest detailed case series published to date included 1,099 hospitalized patients with laboratory-confirmed Covid-19 infection in China, of whom only 25 (2.3%) underwent invasive ventilation and 15 (1.4%) died.^1^ By contrast, 28.1% of hospitalized patients with definitive outcomes in this case series have so far required invasive ventilation and 18.5% have died. Given the very high prevalence of disease in New York City and the relative paucity of baseline hospital beds per capita (1.5-2.7 beds per 1,000 in all boroughs except Manhattan), admission thresholds may be higher in New York City than in China (4.2 beds per 1,000).^21,22^ Moreover, in the series by Guan et al, only a quarter of the hospitalized patients had any chronic comorbidity, whereas in our series 71.9% of hospitalized patients had at least one chronic disease.

In fact, outcomes in the majority of reports are similar to ours. A commentary by the Chinese Center for Disease Control and Prevention described outcomes for 72,314 cases, of which 14% were severe (similar to hospitalized patients in our series) and 5% critical with respiratory or multiorgan failure (similar to those with critical illness in our series).^23^ Among critical cases, mortality was 49%; ours is 45% to date. This is also similar to the typical mortality rate from acute respiratory distress syndrome (ARDS) of about 35-45%.^24,25^ Finally, our results are also consistent with a recent national case series reported by the US CDC that found that 457 of 1,037 (44%) hospitalized patients required ICU admission, and that three quarters had at least one chronic condition.^5^

The risk factors we identified for hospitalization in Covid-19 are largely similar to those associated with any type of severe disease requiring hospitalization or ICU level care, though we were surprised that cancer and chronic pulmonary disease did not feature more prominently in the risk models.^26^ Moreover, the demographic distribution of hospitalized patients is also similar to other acute respiratory infections. For instance, while advanced age was by far the most important predictor of hospitalization and an important predictor of severe outcomes (as it is for most illnesses), 54% of hospitalized patients were younger than 65 years. This is typical of the hospitalization pattern in viral respiratory disease. Studies of influenza hospitalizations in the United States have found that people younger than 65 years account for 53-57% of influenza-related hospitalizations.^27,28^ While men made up a grossly disproportionate number of both hospitalizations and critical illness, this difference was attenuated by multivariable adjustment for comorbidities such that gender was no longer one of the most prominent risk variables.

Surprisingly, though some have speculated that high rates of smoking in China explained some of the morbidity in those patients, we did not find smoking status to be associated with increased risk of hospitalization or critical illness. This is consistent with a handful of other studies that have previously shown a lack of association of smoking with pulmonary disease-associated ARDS (i.e. from pneumonia), as compared with non-pulmonary sepsis-associated ARDS.^29^

More striking were our findings about the importance of inflammatory markers in distinguishing future critical from non-critical illness. Among these, early elevations in c-reactive protein and d-dimer had the strongest association with mechanical ventilation or mortality. Hyperinflammatory states are well described in severe sepsis;^30^ however, the degree to which Covid-19 related inflammation is similar to or different than that typically found in sepsis is unknown. Some emerging case reports suggest that patients with critical Covid-19 disease are developing complications from hypercoagulability,^8^ including both pulmonary emboli^31^ and microscopic thrombi.^32^ In this regard it is notable that the chronic condition with the strongest association with critical illness was obesity, with a substantially higher odds ratio than any cardiovascular or pulmonary disease. Obesity is well-recognized to be a pro-inflammatory condition.^33,34^ Finally, we noted that early (relatively mild) elevation in procalcitonin was a powerful splitter in the classification tree, although Covid-19 appears to be characterized by low procalcitonin levels in general. While many patients with elevated procalcitonin and critical illness were treated with antibiotics, it remains unclear whether these patients actually had bacterial disease or whether the elevation in procalcitonin was another manifestation of a general hyperinflammatory state.

This study includes several limitations. We did not have access to symptom duration which is an important predictor of hospitalization: patients rarely require hospitalization with less than a week of symptoms. However, this limitation should not affect the demographic and clinical characteristics of those requiring admission and having severe deterioration. Importantly, as we are still early in our epidemic, many patients do not yet have final outcomes established, though the sample size of those who do is still more substantial than any prior study of associations with outcomes. Our patients were all from a single geographic region, treated within a single health system; factors associated with poor outcomes may differ elsewhere, though our patient population is very diverse. We did not have inflammatory markers available for non-hospitalized patients; it is possible that these would have been strong predictors for hospitalization risk as well if available. Finally, a standardized admission laboratory protocol was only established about two weeks into the epidemic, resulting in missing laboratory data for earlier patients, especially those who were less acutely ill.

Overall, we find that age and comorbidities are powerful predictors of requiring hospitalization rather than outpatient care; however, degree of oxygen impairment and markers of inflammation are strongest predictors of poor outcomes during hospitalization. Clinicians should consider routinely obtaining inflammatory markers during hospitalizations for Covid-19.

## Data Availability

Individual level data are not available for this study. For aggregate data please contact the corresponding author.

## Acknowledgements

The authors thank Brian Bosworth, Steven Chatfield, Robert Grossman, Thomas Doonan and Daniel Widawsky for operational and technical support. We also thank the thousands of NYU Langone Health employees who have cared for these patients.

